# pyPatlak: Open-Source Voxel-Wise Patlak Analysis of Dynamic PET Data

**DOI:** 10.1101/2025.09.16.25335861

**Authors:** Alessia Artesani

**Affiliations:** Department of Biomedical Sciences, Humanitas University, via Rita Levi Montalcini 4, 20072 Pieve Emanuele, Milan, Italy; IRCCS Humanitas Research Hospital, via Manzoni 56, 20089 Rozzano, Milan, Italy

## Abstract

**Background:** The Patlak graphical analysis is a widely used method for quantifying irreversible tracer uptake in dynamic Positron Emission Tomography (PET) studies, providing key kinetic parameters such as the net influx rate (*K*_*i*_) and the total distribution volume (*V*_*d*_). Currently, this analysis is primarily performed using proprietary software or vendor-specific workstations, which can limit its accessibility, flexibility, and reproducibility. This work presents pyPatlak, an open-source, Python-based, platform-independent tool that performs Patlak modelling directly from dynamic PET DICOM data, offering an accessible alternative to proprietary software.

**Methods:** The pyPatlak script processes dynamic PET data in DICOM format to generate 3D parametric images of *K*_*i*_ and *V*_*d*_. Key features include the normalization of a population-based input function (PBIF) with a patient-specific arterial input function (AIF), and optional correction for the partial volume effect (PVE). The script performs a voxel-by-voxel linear regression to derive the kinetic parameters. To validate the script, we compared the *K*_*i*_ and *V*_*d*_ values generated by pyPatlak with those obtained from a commercial workstation’s direct Patlak analysis. This validation was performed on 21 patients by segmenting seven organs of interest and comparing the values of the kinetic parameters.

**Results:** PyPatlak showed a good agreement with the reference direct Patlak reconstruction. Correlation analysis demonstrated strong linear relationships (*K*_*i*_: R = 0.91, *V*_*d*_: R = 0.93), and Lin’s concordance coefficients confirmed high agreement (*K*_*i*_: 0.89, *V*_*d*_: 0.91). Bland–Altman analysis indicated that observed differences were minimal and clinically negligible. Mean biases were approximately –0.03 ml/min/100ml for *K*_*i*_ and +2.2 units for *V*_*d*_. Equivalence testing further confirmed that all differences fell within predefined clinically acceptable limits, despite being statistically significant in Wilcoxon signed-rank tests.

**Conclusion:** pyPatlak offers a flexible, reproducible, and transparent alternative to proprietary software for Patlak analysis. Its open-source nature and compatibility with standard DICOM data make it a valuable tool for researchers, promoting greater accessibility and standardization of kinetic modelling in PET imaging.

## I. INTRODUCTION

Dynamic Positron Emission Tomography (PET) provides crucial information on physiological and pathophysiological processes by measuring tracer uptake over time [1]. To extract meaningful quantitative information, kinetic models are applied to time-activity curves (TAC) from different tissues [2]. Patlak graphical analysis is a linear graphical method widely used for two-tissue compartments with irreversible uptake, such as ^18^F-fluorodeoxyglucose (FDG) [3][4]. This method allows for the estimation of two key parameters: the net influx rate (*K*_*i*_, Patlak slope) and the total distribution volume (*V*_*d*_, Patlak intercept), which are essential to understand metabolic activity [5][6].

Despite its utility, the application of Patlak analysis in research is often constrained. The required software is typically proprietary and expensive (for example, PMod, OsiriX, MATLAB with specific toolboxes) or integrated into vendor-specific workstations (e.g., Siemens Healthineers) [7] [8] [9] [10]. Relying on closed-source solutions can create significant barriers, including high costs, limited customization, and lack of transparency. These factors can hinder collaboration and reproducibility.

To overcome these challenges, we developed pyPatlak, an open-source Python script designed for voxel-wise Patlak analysis. This paper describes the features of the pyPatlak script and presents a validation study comparing its results with those obtained from a standard commercial workstation. The code is freely available on GitHub [11].

## II. MATERIALS AND METHODS

### A. The pyPatlak Script

pyPatlak is a command-line Python script designed to perform a voxel-wise Patlak graphical analysis on dynamic PET data. The main module, `patlak.py`, contains the core functionality, including DICOM parsing, data conversion, and the Patlak calculation itself. The script is designed to be easily installable and executable via a `setup.py` file.

The pyPatlak workflow is designed to be fully automated, starting from raw DICOM files and ending with parametric NIfTI maps. The principal steps are as follows:

#### (1) Input Parsing

The script takes four essential arguments: the directory containing the dynamic PET DICOM files, the path to a NIfTI mask of the patient-specific arterial input function (AIF), the desired output directory, and a patient ID for file naming. Additional arguments include the path to a population-based input function (PBIF) CSV file, the path to the `dcm2niix` executable, and a recovery factor to correct for the partial volume effect and to scale the mean values of the AIF mask.

#### (2) DICOM-to-NIfTI conversion

The script uses the external command-line tool `dcm2niix` to convert the dynamic series of DICOM images into single NIfTI files, one per frame [12].

#### (3) Patlak Graphical Analysis

The script applies a voxel-wise linear regression according to the Patlak model to derive parametric images of *K*_*i*_ and *V*_*d*_. The results are saved as two separate NIfTI files.

### B. Input Function Management

pyPatlak adopts a hybrid strategy for input function handling that combines the practicality of a population-based input function (PBIF) with patient-specific information. The FDG PBIF, provided as a CSV file, was generated at our institution using a Bayesian optimization procedure on a reference dataset [13]. It is particularly useful for short dynamic FDG PET acquisitions (about 20 minutes), as it avoids the need for full 60-minute protocols or invasive arterial sampling. Alternatively, if the user has their own input function available, it can be supplied directly at the command line, replacing the default PBIF.

To account for inter-patient variability, the PBIF is scaled using the tracer concentration measured in the patient’s artery. The user provides an AIF mask, from which mean activity is extracted for each frame. An optional recovery factor can be applied to correct for partial volume effects when scanner-specific calibration is available. The normalized PBIF is then obtained by rescaling according to the ratio of the patient’s arterial activity to the population curve. Finally, the script checks the alignment between the user-provided AIF mask and the PET images. If orientations differ, axes are flipped automatically to ensure proper registration and avoid misalignment.

### C. Voxel-wise Patlak Analysis

The core of the analysis is a voxel-by-voxel linear regression based on the Patlak model. The script performs the following steps for each voxel. For each time point (*t*) in the dynamic acquisition, the script calculates the two key variables for the Patlak plot [14], which are:

- Independent Variable (X): The time-integrated corrected PBIF (*C*_*p*_(*t*))divided by the corrected PBIF at time *t*, denoted as

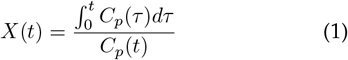
- Dependent Variable (Y): The tissue concentration at time t (*C*_*T*_(*t*))divided by the corrected PBIF at time *t*, denoted as

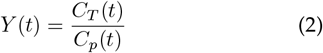

The least-squares linear regression is performed on the data points (*X*_*j*_, *Y*_*j*_)for each voxel, where *j* represents a frame in the dynamic acquisition. The slope of the regression line corresponds to the net influx rate (*K*_*i*_), while the y-intercept is related to the total distribution volume (*V*_*d*_). In our implementation, *K*_*i*_ is reported in ml/min/100ml unit, while *V*_*d*_ is a dimensionless parameter expressing the ratio of plasma volume required to contain the same amount of unmetabolized tracer as 1 mL of tissue. The resulting *K*_*i*_ and *V*_*d*_ values for each voxel are stored in two separate 3D arrays. These arrays are then saved as standard NIfTI files (`Ki map.nii` and `Vd map.nii`) with the same affine transformation as the original PET images, ensuring correct spatial representation. The script sets any negative values of parametric images to zero, as these values are considered unphysiological.

### D. Validation Study

To validate `pyPatlak`, we conducted a retrospective analysis using dynamic FDG-PET/CT scans acquired as part of a previously approved imaging study (IRCCS Humanitas Research Hospital, Ethics Committee Approval ID: NUNC-001-2024, Prot. No. 351/24). The scans were originally collected for research purposes and were subsequently repurposed for technical validation of the algorithm.

The dataset included 21 patients with suspected or confirmed solid tumours. Each patient received a weight-based FDG injection (4.8 MBq/kg), followed by a 50-minute uptake period. A dynamic whole-body (dWB) PET/CT scan was then acquired using continuous bed motion (20 minutes total, 4 passes of 5 minutes each). A low-dose CT scan was acquired first for attenuation correction and anatomical localization.

PET images were reconstructed using an OSEM algorithm (4 iterations, 5 subsets), incorporating time-of-flight and resolution modelling. Corrections were applied for attenuation, scatter, random coincidences, dead-time, and radioactive decay. No post-reconstruction filtering was used. The resulting images comprised 335 planes (440 × 400 voxels) with a voxel size of 1.65 × 1.65 × 3 mm^3^.

All PET data were fully anonymized prior to analysis. An image-derived arterial input function (AIF) mask was generated by manually drawing a circular region of interest (approximately 15 mm in diameter) in the descending aorta and saved as a NIfTI file.

Dynamic PET data were analysed using both the vendor’s direct Patlak reconstruction (reference method) and the `pyPatlak` script. In the reference workflow, the same PBIF was uploaded, and the software automatically extracted a patient-specific AIF from the co-registered CT to scale the PBIF. Parametric images were then generated directly from the raw PET sinograms. In contrast, `pyPatlak` operates on reconstructed dynamic PET images, applying voxel-wise linear regression to indirectly generate Patlak parametric maps.

### E. Statistical Analysis

Comparison was made on values extracted from organ delineation, performed automatically using the TotalSegmentator tool [15] that provides direct segmentation of major organs from the co-registered CT. These segmentations defined the regions of interest (ROIs) used to extract mean, standard deviation, and maximum values of the *K*_*i*_ and *V*_*d*_ maps generated by pyPatlak and by the direct Patlak (reference) reconstruction. For each organ, the mean, the maximum values and the standard deviation are provided.

The comparison between methods was carried out using the Wilcoxon signed-rank test on paired ROI values of *K*_*i*_ and *V*_*d*_. Agreement was quantified by Bland–Altman analysis (bias and 95% limits of agreement) and by Lin’s concordance correlation coefficient (CCC). Equivalence was assessed using the Two One-Sided Tests (TOST) procedure with predefined bounds of ± 0.10 ml/min/100ml for *K*_*i*_ and ± 5 units for *V*_*d*_. These thresholds correspond to approximately 5-10% of typical values, and are in line with reproducibility estimates from recent dynamic whole-body FDG PET/CT studies [16], which observed test–retest variation of *<*19% for multiparametric parameters in many normal tissues. Differences within these limits are therefore considered clinically negligible.

## III. RESULTS

The validation analysis included 21 patients and seven anatomical regions. Table I reports the average *K*_*i*_ and *V*_*d*_ values across organs of interest, together with the mean differences between pyPatlak and the direct Patlak (reference). Differences in *K*_*i*_ ranged from –0.09 to +0.09 ml/min/100ml, while for *V*_*d*_ they varied between –5 and +11 units. For example, in the liver the mean *K*_*i*_ was 0.35 ml/min/100ml with pyPatlak compared to 0.44 ml/min/100ml with the reference (Δ= −0.09), whereas in the kidney the corresponding values were 0.53 and 0.54 ml/min/100ml (Δ= −0.01). Correlations across organs were consistently high (R = 0.85–0.99), with the strongest agreement in kidneys and liver and slightly lower in gallbladder and spleen.

**Table I.**
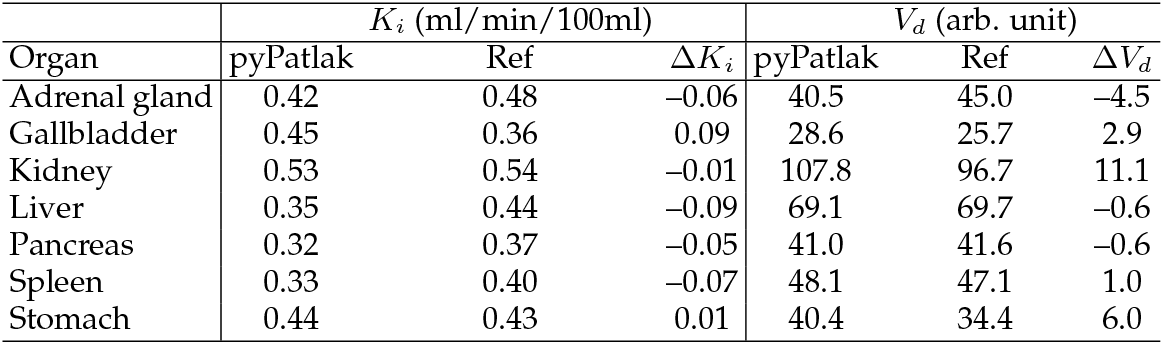
Organ-wise comparison of *K*_*i*_ and *V*_*d*_ between pyPatlak and direct Patlak (reference).

Across all ROIs, the mean bias for *K*_*i*_ was –0.03 ml/min/100ml (95% CI: –0.04 to –0.01), with 95% limits of agreement from –0.20 to +0.15 ml/min/100ml. For *V*_*d*_, the mean bias was +2.2 units (95% CI: +0.5 to +4.3), with limits of agreement from –21.8 to +26.2. Maximum values were also consistent between methods, with *K*_*i*_ peaks around 0.5–0.6 ml/min/100ml and *V*_*d*_ values up to 100 units. These small systematic differences indicate that the two methods provide highly comparable results within the expected variability of parametric PET images.

Linear regression analysis confirmed strong global correlations between methods (Figure 1A,B), with R = 0.91 for *K*_*i*_ and R = 0.93 for *V*_*d*_. Lin’s concordance correlation coefficient further supported this agreement (*K*_*i*_: CCC = 0.89; *V*_*d*_: CCC = 0.91). Bland–Altman plots (Figure 1C,D) showed differences symmetrically distributed around zero without systematic bias. Using predefined equivalence bounds of ±0.10 ml/min/100ml for *K*_*i*_ and ±5 units for *V*_*d*_, the Two One-Sided Tests (TOST) procedure confirmed equivalence for both parameters. This demonstrates that although Wilcoxon signed-rank test detected statistically significant differences (*K*_*i*_: p *<* 0.001; *V*_*d*_: p = 0.034), these remain negligible in magnitude and clinically irrelevant. Taken together, the results show that pyPatlak produces *K*_*i*_ and *V*_*d*_ values in close agreement with the direct Patlak (reference) implementation, supporting its reliability as an open-source alternative for voxel-wise Patlak analysis in dynamic PET.

**Figure 1.**
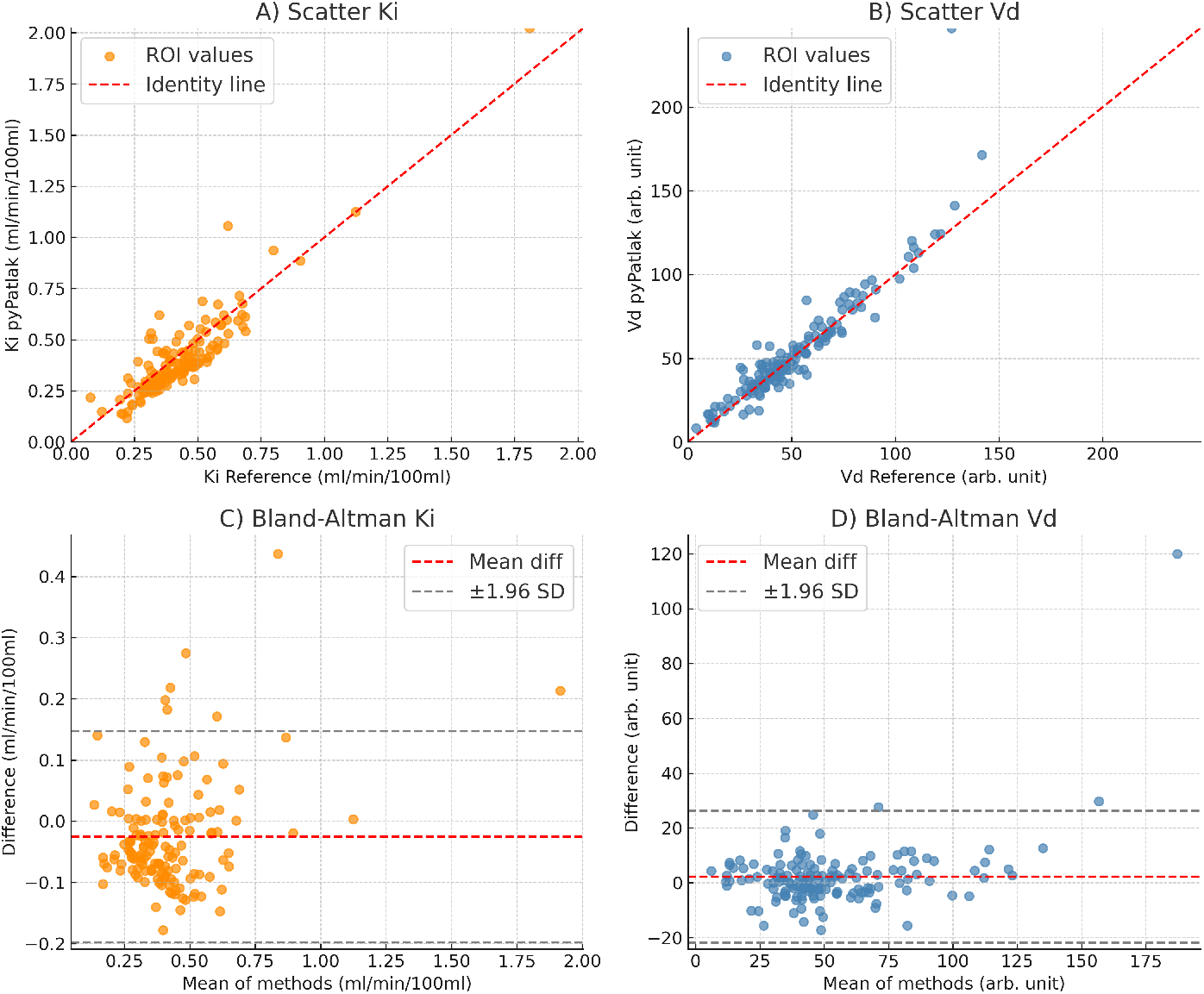
Comparison between pyPatlak and direct Patlak (reference) analysis. (A) Scatter plot of mean *K*_*i*_ values across all ROIs, showing strong correlation (R = 0.92, p *<* 0.001). (B) Scatter plot of mean *V*_*d*_ values, with excellent correlation (R = 0.97, p *<* 0.001). (C) Bland–Altman plot of *K*_*i*_ values, with mean difference (red line) and limits of agreement (±1.96 SD, grey lines), demonstrating no systematic bias. (D) Bland–Altman plot of *V*_*d*_ values, confirming good agreement without systematic deviation. *K*_*i*_ values are expressed in ml/min/100ml and *V*_*d*_ values in arbitrary units.

## IV. CONCLUSION

We have developed and validated pyPatlak, an opensource Python script for voxel-wise Patlak analysis of dynamic PET data. The tool provides a practical and transparent alternative to proprietary implementations, enabling quantitative PET analysis that is accessible and reproducible. Validation against a reference direct Patlak reconstruction confirmed its accuracy and reliability, with differences that were statistically detectable but clinically negligible. By fostering transparency and interoperability, pyPatlak can support collaborative and standardized studies in quantitative PET imaging and facilitate its broader adoption in research and clinical applications.

## Data Availability

The code used in this study is publicly available. The PET imaging data used for validation are not publicly available due to patient privacy considerations.

https://github.com/alessiaartesani/PyPatlak

## Data Availability

https://github.com/alessiaartesani/PyPatlak

## AUTHOR APPROVAL

All authors have seen and approved the manuscript.

## ETHICS APPROVAL

This study involved the technical validation of an open-source computational method using previously acquired PET imaging data. All data were fully anonymized and analyzed retrospectively. The data collection and use were approved by the Ethics Committee of IRCCS Humanitas Research Hospital (Approval ID: NUNC-001-2024, Prot. No. 351/24), in accordance with the principles of the Declaration of Helsinki.

## INFORMED CONSENT

Informed consent was obtained at the time of acquisition, and all data were fully anonymized prior to analysis.

## DATA AVAILABILITY STATEMENT

The code used in this study is publicly available at [11]. The PET imaging data used for validation are not publicly available due to patient privacy considerations.

